# Humoral Response to BNT162b2 mRNA Covid19 Vaccine in Peritoneal and Hemodialysis Patients: a Comparative Study

**DOI:** 10.1101/2021.06.14.21258113

**Authors:** Rui Duarte, Marisa Roldão, Cátia Figueiredo, Francisco Ferrer, Hernâni Gonçalves, Ivan Luz, Flora Sofia, Karina Lopes

## Abstract

**Introduction:** Generalized immunization against COVID19 has become the cornerstone in prevention of severe acute respiratory syndrome associated with this pandemic. Maintenance dialysis patients (MDP) are at higher risk of both exposure and mortality from the disease. Efficacy and security of BNT162b2 vaccine is well documented for the general population, but not in MDP, particularly in peritoneal dialysis (PD) patients. This study aims to compare humoral response between hemodialysis (HD) and PD patients.

**Materials and Methods:** Observational prospective study including MDP on HD or PD program from a Portuguese middle-sized Nephrology Center, who received BNT162b2 vaccine. Specific anti-Spike IgG was measured as arbitrary units per milliliter (AU/mL) on two separate occasions: 3 weeks after the first dose and 3 weeks after the second. The two modality groups were compared both for absolute value and number of non-responders (NR) after both inoculations. Demographic data was also compared.

**Results:** Of 73 patients enrolled, 67 were eligible for the final study: 42 HD and 25 PD patients. PD group developed significantly higher antibody titers in both inoculations: first dose with Med 5.44 vs 0.99 (p<0.01) and second dose with Med 170.43 vs 65.81 (p<0.01). HD status was associated with NR after the first dose (Phi=0.383; p<0.01), but not after the second (p=0.08). Age, Charlson comorbidity index and dialysis vintage were lower in the PD group (p<0.01; p=0.02; p<0.01, respectively).

**Conclusion:** This study demonstrated a better humoral response to immunization with BNT162b2 in PD patients, when compared to HD patients, after each of the two recommended inoculations. Both groups showed substantial humoral response after just one dose of the vaccine. Older age and higher comorbidity burden may explain the relative immunogenicity deficit, probably in a superior degree comparing with age matched healthy population.

**What is already known about this subject:**

- Maintenance hemodialysis patients have lower humoral response to BNT162b2 mRNA COVID19 vaccine when compared to the general population;
- Maintenance dialysis patients are at high risk of exposure to COVID19 in addition to a more severe course of the disease;

**What this study adds:**

- Maintenance peritoneal dialysis patients have better humoral response with BNT162b2 when compared to those on hemodialysis;
- There is a substantial humoral response after a single dose of the vaccine for both hemodialysis (50%) and peritoneal dialysis (88%) patients.

**What impact this may have on clinical practice or policy:**

- Protocols for follow-up measures, including extra inoculations, might have to be considered for hemodialysis patients;
- Peritoneal dialysis patients should be promptly immunized in all centers, rejecting constraints regarding lower effectiveness or yield.

## Introduction

Worldwide vaccination against coronavirus 2019 (COVID19) has become the cornerstone in prevention of Severe acute respiratory syndrome Coronavirus 2 (Sars-CoV-2) associated with this pandemic. Several vaccines with different acting mechanism have been developed, namely BNT162b2 (Pfizer BioNTech)^1^ and mRNA-1273 (Moderna)^2^ – mRNA-based vaccines; ChAdOx1 nCov-19 (AstraZeneca)^3^ and Ad26.COV2.S (Johnson&Johnson/Janssen)^4^ – recombinant adenovirus vectors encoding Sars-CoV-2 spike glycoprotein, among others.

The need for mandatory regular contact with health care services in maintenance dialysis patients (MDP), coupled with worse disease severity and increased mortality^5-7^, establish MDP as a high-risk population. Following this assessment and international recommendations, the Portuguese government implemented vaccination for MDP early in the immunization plan, starting on February 2021.

For a long time, antiviral immunogenicity in MDP patients has been a known issue, particularly concerning antiviral vaccination, as illustrated by hepatitis B with only 50 to 60% of hemodialysis (HD) patients exhibiting humoral response in the first studies when compared to over 90% in healthy individuals^8,9^. To minimize this problem, new protocols were established involving follow-up antibody measurements, adjuvant and new vaccines development and repeated inoculations, improving seroconversion in HD patients to 80%^10^. Data on peritoneal dialysis, however, is sparser.

The efficacy studies for COVID19 vaccines described above did not include MDP. Humoral response to BNT162b2 in HD patients has been a target in multiple studies^11^, revealing lower titers compared to general population, however, there is still no data on PD patients. Age has also been suggested as a risk factor for worse humoral response^11^. This study aims to compare humoral response to the BNT 162b2 in our Nephrology center between PD and HD patients.

## Materials and Methods

The study design consisted of an observational prospective study. It included a group of forty-six HD and twenty-seven PD patients who were scheduled to receive two doses of BNT 162b2 in a three-week interval, in accordance with pharmaceutical guidelines for its administration, between February and March 2021.

The assessment of humoral vaccination response was done as part of the internal policy of the center ‘s contingency protocol and informed consent of each patient was obtained regarding the use and access to these analytical results, as well as the remaining and sociodemographic information for scientific research. Blood collection and analysis was made at two distinct phases: 1) Three weeks after the inoculation of the first dose and 2) Three weeks after the second dose. Because the recommended dosing interval is three weeks, the first collection was therefore coincidentally performed with the second dose administration.

Humoral response as IgG anti-Spike for COVID19 was measured in addition to IgM both for anti-Spike and anti-Nucleocapside for tracking possible contacts, even if asymptomatic. Titers were measured as arbitrary units per milliliter (AU/mL) using chemiluminescence. Response was considered significant for titers superior to 1 AU/mL.

Inclusion criteria included capacity to understand and provide informed consent and who did not show a significant increase in specific IgM anti-N for COVID19 during follow-up.

We hypothesized that both humoral response, as measured by IgG anti-Spike, and response rate would be higher in the PD group. As primary endpoints we established the comparison between the median of achieved titers to both the first and second dose and between rate of non-responders (NR) for each modality. Secondary endpoints focused on comparison of demographic data including age, sex, diabetes, Charlson comorbidity index (CCI) and dialysis vintage.

## Statistical analysis

Statistical analysis was carried out using Microsoft Excel 2016 and IBM SPSS Statistics 25 software.

Study variables, divided in demographic and humoral, are summarized in table 1. Descriptive analysis was performed using frequencies and percentages for categorical variables, whereas continuous variables are presented using means with standard deviations, if normal distributed, or medians with interquartile range, for skewed distribution.

**Table 1:** Humoral and demographic variables in the study.

| Continuous | Categorical |
| --- | --- |
| **Humoral response associated variables** |  |
| IgG anti-Spike S1 levels 1<sup>st</sup> dose (AU/mL) | 1<sup>st</sup> dose NR (<1 AU/mL) |
| IgG anti-Spike S1 levels 2<sup>nd</sup> dose (AU/mL) | 2<sup>nd</sup> dose NR (<1 AU/mL) |
| **Demographic** |  |
| Age | Sex |
| CCI | Diabetes |
| Dialysis vintage |  |

NR: Non-responders; AU/mL: Arbitrary units per milliliter; CCI: Charlson comorbidity index

Demographic data, anti-spike IgG levels and NR rate were compared between both modality groups. Differences in continuous variables between groups were analyzed with Student ‘s *t*-test for means (parametric) and Mann-Whitney *U* test for medians (non-parametric). The response rate was also analyzed between modalities using Fisher ‘s exact test and Phi correlation coefficient. Pearson and Spearman correlation tests between antibody levels and continuous variables were also performed, if normal distributed or not, respectively.

## Results

Of the forty-six HD patients enrolled, forty-two were eligible for the study: 1) two patients discontinued dialysis before the administration of the second dose; 2) one patient died from an unrelated cause and 3) despite asymptomatic one patient showed a significant increase in IgM titer raising the possibility of contact with the virus. Similarly, two patients from the PD group showed significant increase in specific IgM and were considered to have asymptomatic infection, resulting in twenty-five PD patients being enrolled into the statistical analysis, a total of sixty-seven MDP.

Demographic data is summarized in table 2, both for descriptive and comparative analysis. PD group was younger (60.5 vs. 75.1; t(65)=5.1; p<0.01), with shorter dialysis vintage (18 vs. 35; U= 348; p= 0.02) and lower CCI (5.2 vs. 7.8; U=229; p<0.01). There was no statistical difference for gender or diabetes between both groups. Humoral response data is summarized in table 3 with boxplot graph representation in figures 1 and 2. IgG titers were higher in PD patients both for the first (5.44 vs. 0.99; U=844; p<0.01) and second doses (170.43 vs. 65.81; U=766; p<0.01). HD was weakly associated with NR for the first dose (Phi 0.383; p<0.01) and showed no association with the second dose (p<0.08) when compared to PD. There were no NR patients in PD group after both doses, an outcome that was observed in 6 patients from the HD group.

**Table 2:**
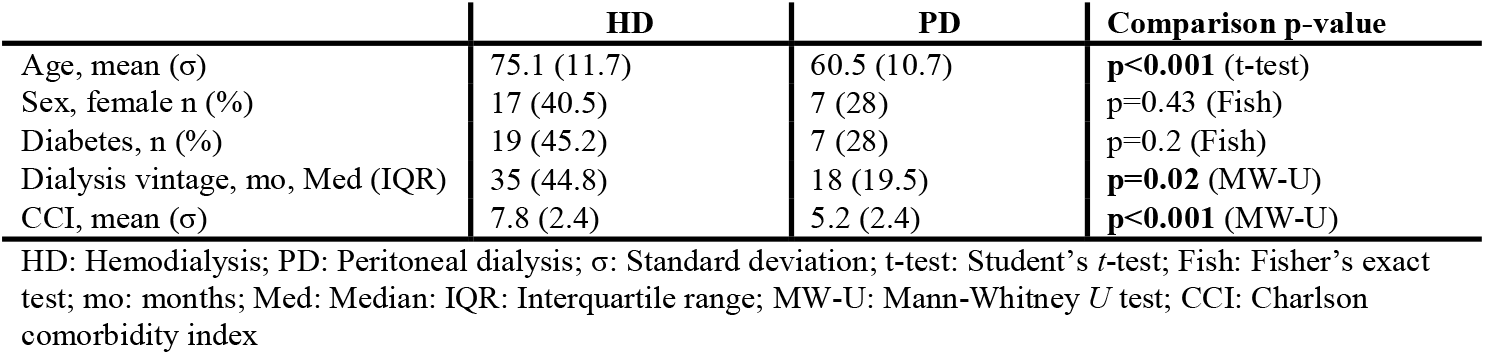
Description and comparison of demographic data by modality.

**Table 3:**
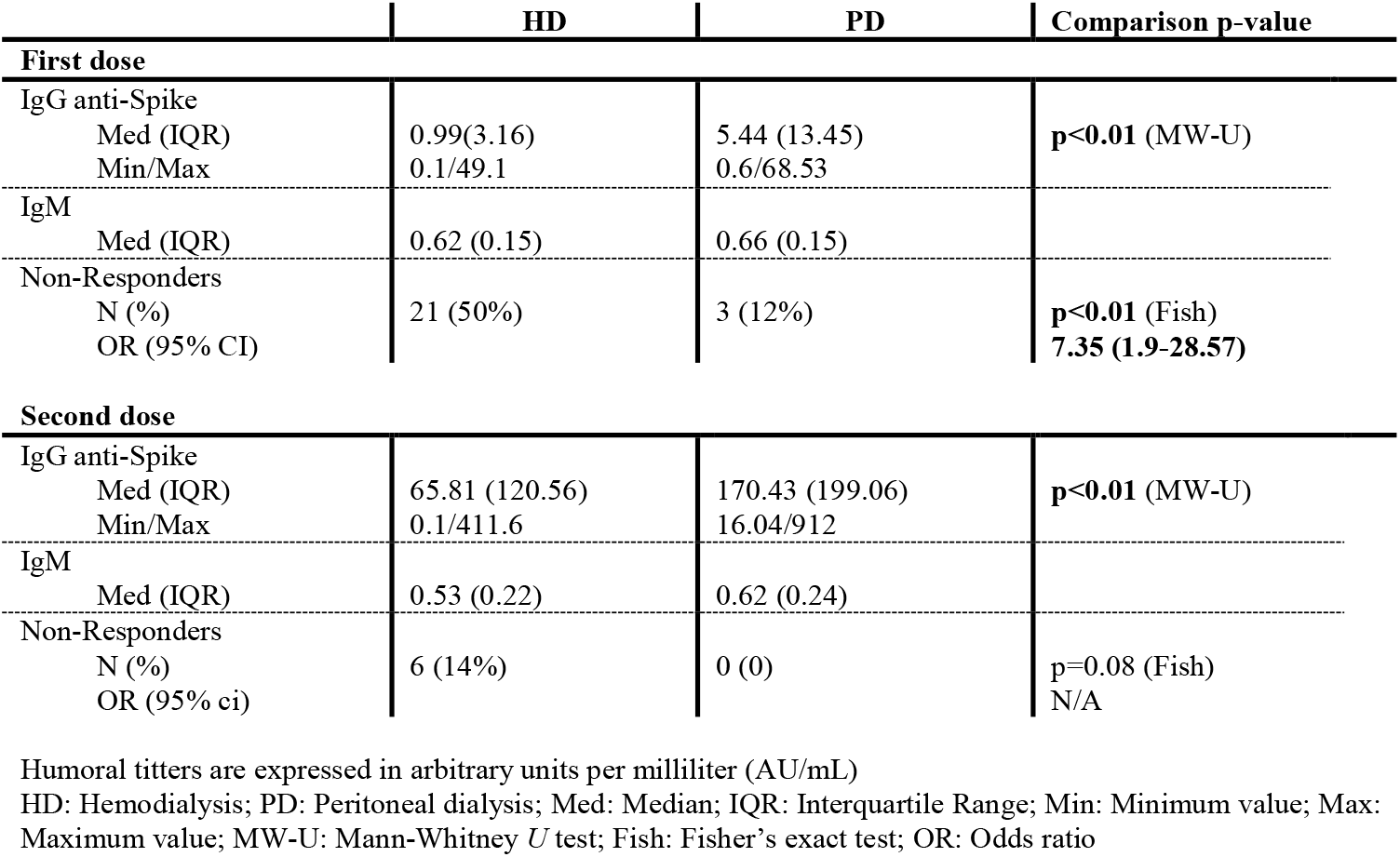
Humoral response titters and rate of non-responders by modality.

**Figure 1:**
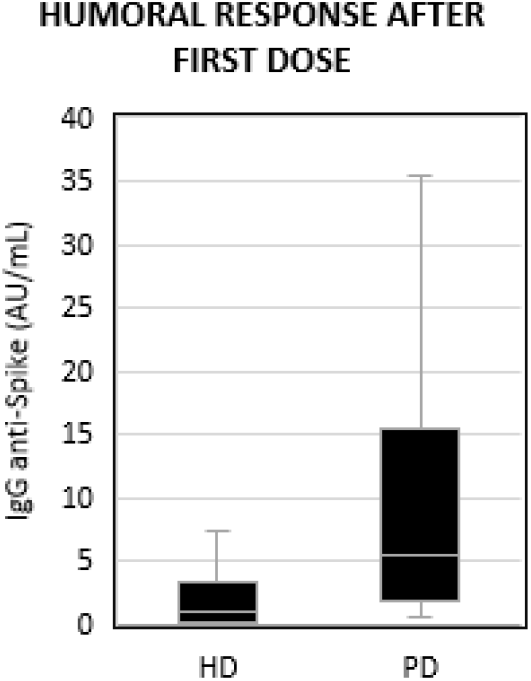
Boxplot graph for first dose humoral response between modalities. AU/mL: Arbitrary units per milliliter; HD: Hemodialysis group; PD: Peritoneal Dialysis group.

**Figure 2:**
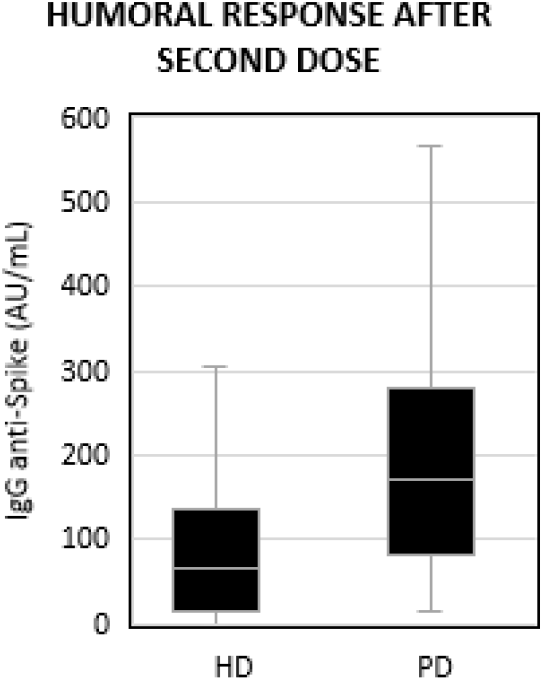
Boxplot graph for second dose humoral response between modalities. AU/mL: Arbitrary units per milliliter; HD: Hemodialysis group; PD: Peritoneal Dialysis group.

## Discussion

Overall, humoral response to BNT162b2 evaluated by IgG anti-Spike titers is significant in MDP with only 9% of the total sample failing to achieve measurable response, all from the HD group. The fact that the difference in NR for the second dose was not statistically different, even though all identified NR were from the HD group, suggests insufficient sample size. Regarding the method to assess antibody immune response (in arbitrary units), it is important to highlight that its laboratory dependence does not allow for external comparison. Our study support that PD patients achieve a significant higher humoral response when compared to the HD counterpart.

The analysis of demographic data favors PD patients as a younger population, with less dialysis vintage and less comorbidity burden (as evaluated by CCI) when compared to HD patients. These differences are expected given the nature, requirements and indications of each modality. Multiple studies have found advanced age to be an independent risk factor for low humoral response to BNT162b2 and it can play a role in our study results.

Approximately half of the patients in the HD group were diabetics, whereas that number dropped to a fifth in the PD group, without a statistically significant difference. It is well known. It is well known that diabetes is associated with an overall immune response dysfunction. However, larger studies may be necessary to establish the role of diabetes in antibody immune response between dialysis modalities.

It is worth noting that, during the follow up no patient developed clinically significant COVID-19 disease or was diagnosed with it via nasopharyngeal swab.

## Conclusion

This study suggests that humoral response in MDP is significantly different for BNT 162b2 vaccination depending on modality, with PD patients showing higher titers after both first and second doses when compared to their HD counterpart. Demographic differences between the two groups must be considered, namely disparities in age and comorbidity burden. The influence of said factors in overall humoral response is unknown. Additional studies are, therefore, required to further explore immunogenicity factors and in dialysis population.

The identification and signalization of higher risk subgroups in MDP for insufficient or no response to vaccination is essential, providing institutions an opportunity to develop and establish protective and follow-up protocols in this pandemic.

To the best of our knowledge, this is the first study evaluating humoral response to BNT162b2 vaccine in PD patients and also comparing them with their HD counterpart.

## Data Availability

The authors confirm that the data supporting the findings of this study are available within the article and its supplementary materials. Specific patient data is not available in accordance to data protection.

## Author contributions

Concept and design: Rui Duarte, Francisco Ferrer, Ivan Luz

Acquisition and interpretation of data: All authors

Drafting of the manuscript: Rui Duarte

Intelectual revision of content: Marisa Roldão, Cátia Figueiredo, Ivan Luz

Statistical analysis: Hernani Gonçalves

Language and final revision: Francisco Ferrer

Technical and administrative support: Flora Sofia

Supervision and approval of the article: Karina Lopes

## Conflict of interest disclosures

There are no conflicts of interest for any of the authors.

## Financial support/funding

No funding was used in this study.

---

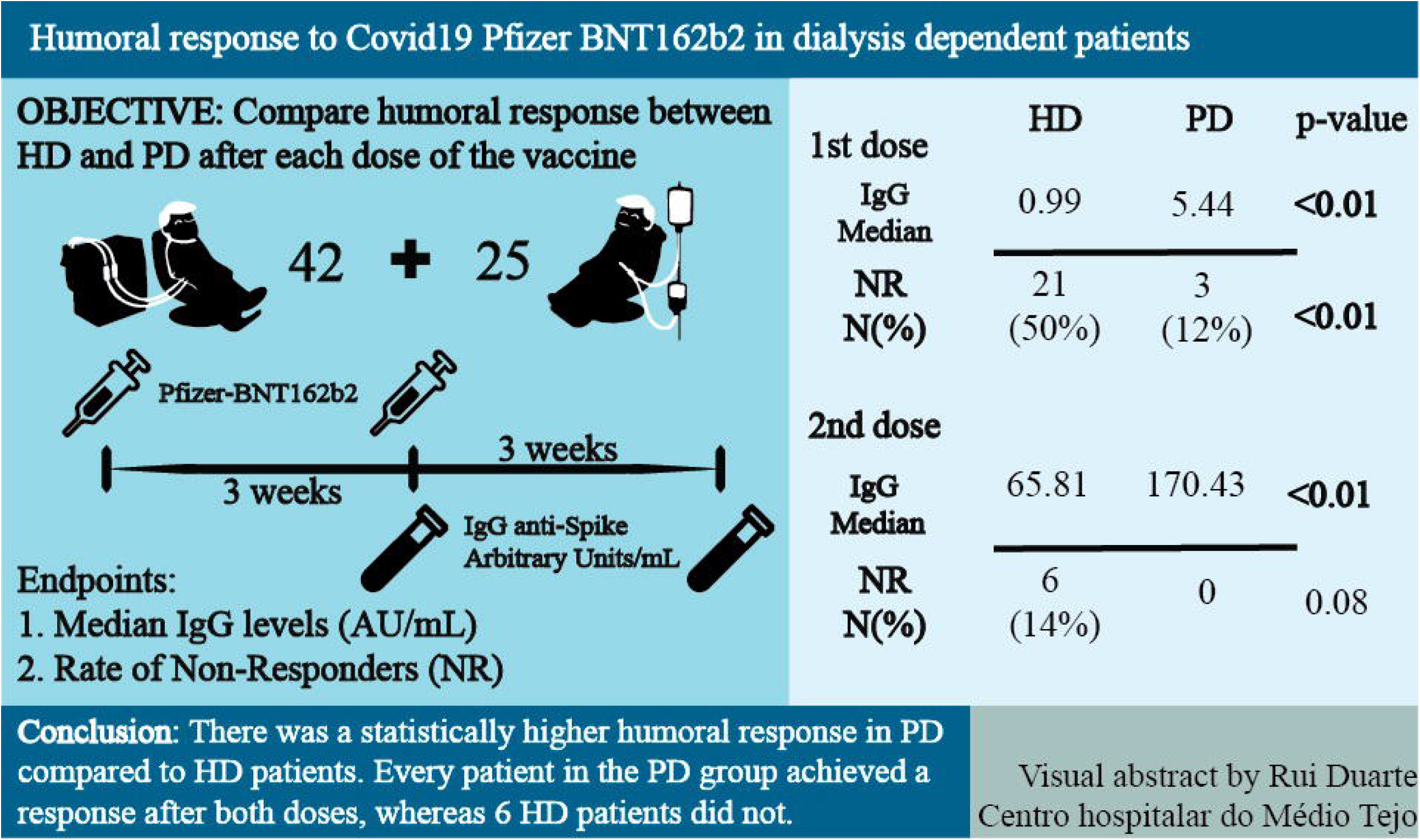

## Notes

### Competing Interest Statement

The authors have declared no competing interest.

### Funding Statement

No funding was received in the making of this study

### Author Declarations

Institutional Ethical Committee of Centro Hospitalar do Medio Tejo

